## Supplemental Materials for "Examining sex differences in neurodevelopmental and psychiatric genetic risk in anxiety and depression"

### Supplemental Text

#### Quality control of NCMH genetic data

DNA samples were collected from venous blood or saliva, where possible. The samples were genotyped using a customised version of the PsychChip in several batches, with rigorous quality control (QC) procedures. QC was performed within batch, as follows: SNPs were aligned to the Haplotype Reference Consortium^1^ data using GenomeHarmoniser^2^, SNPs were removed if they had MAF<0.01, genotyping rate<0.95, or HWE p<10^-6^, and individuals were removed if they had missingness>0.05, sex discrepancy, or were duplicate samples. The data were merged with other samples on the same or equivalent platform and the QC parameters were reapplied, then the samples were imputed using the Michigan Imputation Server (using Eagle v2.4 for phasing, Minimac4 for imputation, and the HRC V1.1 imputation reference panel)^3^. After imputation, dosage data were converted to best guess genotype data using Plink2^4^.

Post-imputation QC filters were genotype probability >0.9 per individual, missingness <0.03, MAF>0.01, HWE>10^-4^, and INFO/r^2^>0.8. Batches were merged, including only overlapping SNPs and excluding ambiguous (CT/AG) variants and SNPs with inconsistent alleles. Family relationships were confirmed using identity-by-descent (IBD) in PLINK. Control individuals were excluded if they had a relative amongst the cases. PCAiR^5^, a package that robustly estimates population structure while taking into account kinship information in the sample, was used to run a principal components analysis (PCA), on an LD-pruned set of common (MAF>0.05) markers and non-European samples were excluded, given that the discovery genome-wide association studies (GWAS) available were of primarily European ancestries^6^. A GWAS of batch was run on unrelated samples and SNPs associated with batch (p<0.01) were excluded. PCAiR was run again on the final set of markers to extract PCs to use as covariates.

#### PRS calculation

Polygenic risk scores were derived using PLINK version 1.9^7^ based on 6 large psychiatric disorder discovery GWAS of primarily European ancestries, with no overlap with the target sample. Data for ADHD PRS for 1 individual from the target sample was excluded as they were included in the published ADHD GWAS. For each discovery GWAS, we selected common (MAF>0.05) variants that overlapped with the target data and performed LD-clumping in PLINK (--clump-kb 500 --clump-r2 0.2) to obtain an independent set of SNPs, while retaining the most significant SNP in each LD (linkage disequilibrium) block. For schizophrenia PRS, we additionally excluded all variants in the extended major histocompatibility complex (MHC), region (chromosome 6, base positions 25–35Mb) to avoid potential bias by extensive LD in this region. PRS were calculated for each individual by summing the number of alleles (weighted by the log of the odds ratio) across the set of SNPs in PLINK (using the command --score). We calculated PRS using 7 different p-value thresholds to select SNPs (p_T_<1, p_T_<0.5, p_T_<0.1, p_T_<0.05, p_T_<0.01, p_T_<0.001, p_T_<0.00001).

For each discovery phenotype, we then performed PCA of the correlation matrix of these 7 PRS and extracted the first PC for analyses, following the PRS-PCA method, an approach that reduces overfitting and has been shown to maintain good power^8^. The sign of the loadings of the PRS variables on the first PCs is arbitrary and therefore PRS-PCs that were negatively correlated with the raw PRS variables were inverted. For each of the 6 discovery phenotypes, the first PRS-PCs explained between 64.2–78.3% of the variation in the different p-value threshold PRS in NCMH. The PRS-PCs were standardised using z-score transformations.

#### Copy number variant (CNV) calling and quality control

CNVs were called in NCMH using PennCNV following standard protocol^9^. CNV calls were merged if the distance separating two CNVs was less than 50% of their combined length. CNV QC was restricted to samples passing SNP-based QC, which included removal of ancestry outliers; see above for details. Individuals were also excluded if they were outliers on any of the following QC metrics generated by PennCNV: Log R Ratio standard deviation (LRR SD) >0.2, waviness factor (WF) >0.03 or <-0.03 and total number of CNVs >100. Following exclusion of these poorly performing samples, CNV QC was performed. CNV calls were filtered out based on the following criteria: size (<100kb), coverage (<20 probes), and PennCNV confidence score (<10).

We identified 54 CNVs impacting on 34 well-defined regions (including duplications and deletions at same loci) implicated in neurodevelopmental disorders (NDs), based on previously published criteria^10^. To be called as genuine ND CNVs, called CNVs underwent further checks that are specific to each locus. These checks included one or more of the following: a) the CNV is required to hit specific genes at a locus, b) exons of a gene may be required to be hit, c) the CNV may be required to be a certain length (e.g. greater than 1MB), and d) the CNV may be required to be longer than a certain percentage of the “critical region”. See published work^10^ for the full list of criteria. Visual inspection of B allele frequency (BAF) and LRR at these loci was performed to confirm these CNVs.

Additional QC steps were then performed on the full set of called CNVs to obtain information on burden of large rare CNVs in each individual. CNVs were further filtered to those that are >500kb in size, and have good probe density (>20kb/probe, calculated as size/probes per CNV). CNVs spanning more than 50% of any of the following regions were excluded: centromeres, telomeres (100kb from ends of chromosomes), known segmental duplications, and immunoglobulin or T cell receptor loci. CNV loci occurring at >1% frequency in the remaining set of samples were removed.

Dichotomous variables were derived for presence of a neurodevelopmental CNV >100kb, any large (>500kb) CNV, as well as large duplications and deletions separately.

#### PGC clinical MDD sample

Age-at-onset of MDD was defined as the age at which individuals self-reported first having symptoms meeting MDD (10,30). Age-at-onset was recoded as missing if it was greater than the age of the participants at interview. Individuals missing information on age-at-onset who were younger than 26 years old at assessment (N=41) were also included in the early onset group. Due to small sample sizes in some of the sub-groups, analyses were restricted to studies with N≥50 total samples and ≥10 individuals on the smaller stratum (2 studies were excluded from the early-onset analysis and 4 studies were excluded from the later onset analysis).

### Supplementary Tables

### Table S1: Clinical and socioeconomic characteristics of males and females with anxiety and depression in NCMH

| **Phenotype (continuous)** | **Males, Mean(SE)** | **Females, Mean(SE)** | **OR(95% CI)** | **P** |
| --- | --- | --- | --- | --- |
| **Age at assessment*** | 42.6 (0.4) | 41.0 (0.3) | 0.99 (0.99-1.00) | 3.5 x 10^-4^ |
| **Age of onset (any psychiatric problems)** | 22.6(0.4) | 20.6(0.2) | 0.99 (0.98-1.00) | 1.4 x 10^-3^ |
| **Age of access to services** | 29.9(0.4) | 26.8(0.3) | 0.98 (0.97-0.99) | 1.2 x 10^-7^ |
| **Age of access to treatment** | 30.1(0.7) | 26.7(0.4) | 0.97 (0.95-0.98) | 1.1 x 10^-7^ |
| **HADS current anxiety symptoms** | 11.6(0.2) | 11.2(0.2) | 0.97 (0.95-1.00) | 0.027 |
| **HADS current depression symptoms** | 9.5(0.3) | 7.7(0.2) | 0.94 (0.92-0.96) | 5.9 x 10^-9^ |
| **Phenotype (dichotomous)** | **Males, N(%)** | **Females, N(%)** | **OR(95% CI)** | **P** |
| **NDs** | 429(29.7) | 424(15.5) | 0.37 (0.32-0.44) | 9.7 x 10^-33^ |
| **OCD** | 146(10.1) | 249(9.1) | 0.87 (0.70-1.07) | 0.19 |
| **PTSD** | 373(25.8) | 528(19.3) | 0.70 (0.60-0.81) | 2.9 x 10^-6^ |
| **Eating disorders** | 44(3.0) | 340(12.4) | 4.41 (3.19-6.08) | 1.7 x 10^-19^ |
| **Substance misuse** | 222(15.4) | 214(7.8) | 0.48 (0.39-0.58) | 3.6 x 10^-13^ |
| **Personality disorder** | 74(5.1) | 215(7.9) | 1.52 (1.16-2.00) | 2.6 x 10^-3^ |
| **Low income** | 667(58.2) | 1270(59.2) | 1.05 (0.91-1.21) | 0.51 |
| **Low education** | 154(12.1) | 209(8.5) | 0.70 (0.56-0.88) | 2.2 x 10^-3^ |
| **NEET** | 437(39.3) | 619(30.0) | 0.67 (0.58-0.79) | 5.5 x 10^-7^ |

* Age at assessment is included as a covariate in all other analyses.

HADS: Hospital Anxiety and Depression Scale; NDs: neurodevelopmental disorders; OCD: obsessive compulsive disorder; PTSD: post-traumatic stress disorder; NEET: not in education, employment or training. Males are coded as 0, females are coded as 1; therefore OR>1 indicates females have a higher likelihood for a given phenotype.

### Table S2: Association of psychiatric polygenic risk scores with anxiety/depression case-control status in NCMH

| **PRS** | **OR(95% CI)** | **P** | **R^2^** |
| --- | --- | --- | --- |
| **ADHD** | 1.28 (1.09–1.51) | 2.7x10^-3^ | 0.011 |
| **ANX** | 1.36 (1.15–1.61) | 4.0x10^-4^ | 0.018 |
| **ASD** | 1.20 (1.02–1.40) | 0.026 | 5.8x10^-3^ |
| **BD** | 1.17 (0.97–1.41) | 0.10 | 4.5x10^-3^ |
| **MDD** | 1.49 (1.25–1.79) | 1.3x10^-5^ | 0.029 |
| **SCZ** | 1.12 (0.93–1.35) | 0.22 | 2.4x10^-3^ |

ADHD: attention deficit hyperactivity disorder; ANX: anxiety disorders; ASD: autism spectrum disorder; BD: bipolar disorder; MDD: major depressive disorder; PRS: polygenic risk score; SCZ: schizophrenia. Controls are coded as 0, cases with anxiety/depression are coded as 1; therefore OR>1 indicates a higher PRS in cases and indicates the effect size per 1 SD of the PRS.

### Table S3: Association of polygenic risk scores for ADHD (primary analysis) and other psychiatric disorders (exploratory analysis) with sex of individuals with anxiety and depression in NCMH, after adjusting for comorbid neurodevelopmental disorders

| **PRS** | **OR(95% CI)** | **P** | **R^2^** |
| --- | --- | --- | --- |
| **ADHD** | 1.03 (0.92-1.16) | 0.57 | 3.2 x 10^-4^ |
| **ANX** | 0.93 (0.83-1.04) | 0.21 | 1.6 x 10^-3^ |
| **ASD** | 1.07 (0.95-1.20) | 0.24 | 1.4 x 10^-3^ |
| **BD** | 1.02 (0.91-1.15) | 0.68 | 1.7 x 10^-4^ |
| **MDD** | 1.01 (0.90-1.13) | 0.88 | 2.5 x 10^-5^ |
| **SCZ** | 0.95 (0.84-1.06) | 0.34 | 9.3 x 10^-4^ |

ADHD: attention deficit hyperactivity disorder; ANX: anxiety disorders; ASD: autism spectrum disorder; BD: bipolar disorder; MDD: major depressive disorder; PRS: polygenic risk score; SCZ: schizophrenia. Males are coded as 0, females are coded as 1; therefore OR>1 indicates a higher PRS in females and indicates the effect size per 1 SD of the PRS.

### Table S4: Association of psychiatric polygenic risk scores with sex of individuals diagnosed with MDD in the PGC replication sample, stratified by age-at-onset

| **Age-at-onset** | **PRS** | **Males** | **Females** | **OR(95% CI)** | **P** | **R^2^** |
| --- | --- | --- | --- | --- | --- | --- |
| **Young (<26 years old)** | **ANX** | 1804 | 4411 | 1.01 (0.96-1.08) | 0.63 | 4.8 x 10^-3^ |
|  | **SCZ** | 1804 | 4411 | 1.00 (0.94-1.06) | 1.00 | 4.9 x 10^-3^ |
| **Older (>25 years old)** | **ANX** | 2100 | 3752 | 0.93 (0.88-0.99) | 0.017 | 4.7 x 10^-3^ |
|  | **SCZ** | 2100 | 3752 | 1.00 (0.94-1.05) | 0.86 | 2.2 x 10^-3^ |

ADHD: attention deficit hyperactivity disorder; ANX: anxiety disorders; MDD: major depressive disorder; SCZ: schizophrenia. Males are coded as 0, females are coded as 1.

### Supplementary Figures

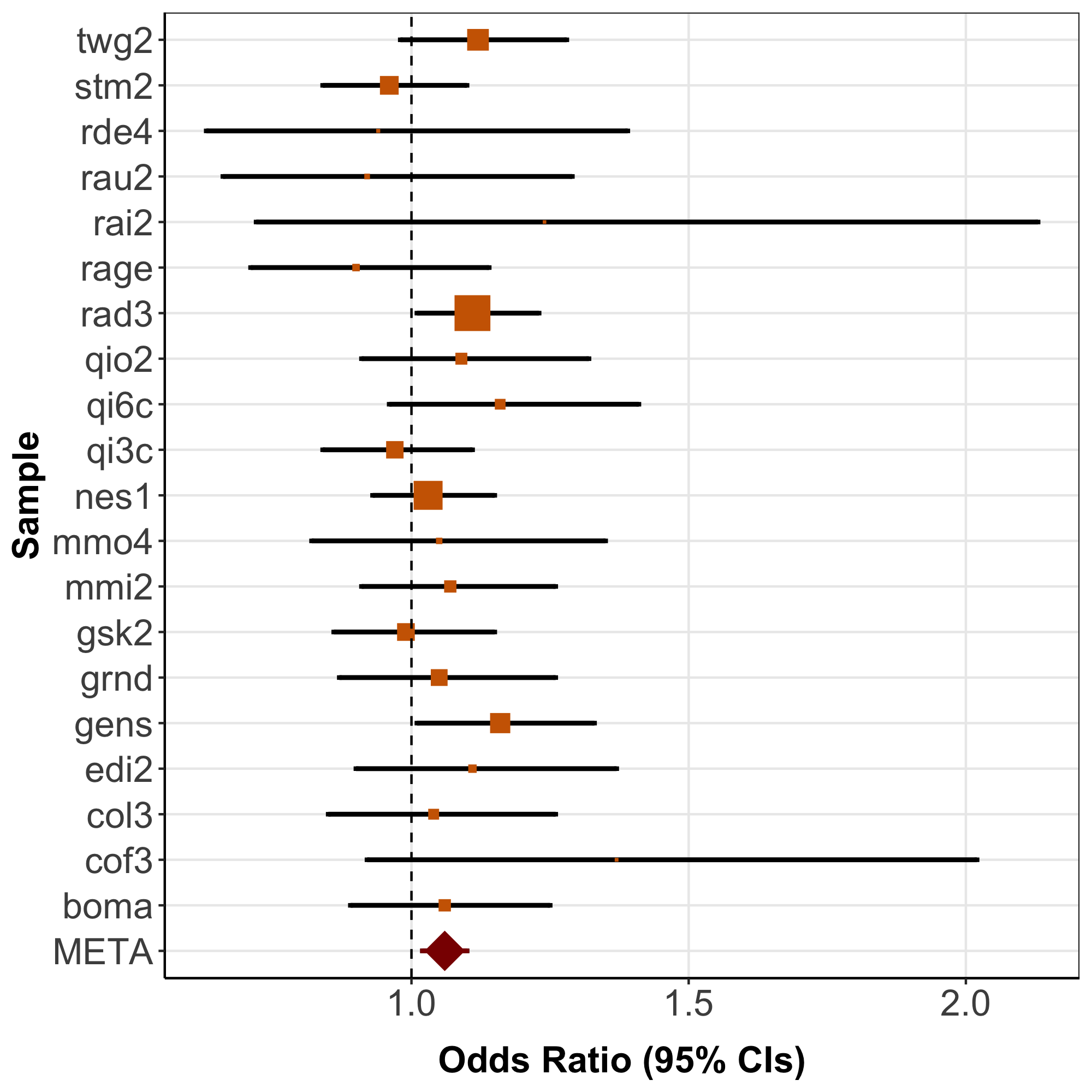

### Figure S1. Forest plot of meta-analysis results for 20 PGC studies for the association between polygenic risk scores for ADHD with sex in individuals diagnosed with major depressive disorder (females coded as 1 and males coded as 0).

**Major Depressive Disorder Working Group of the Psychiatric Genomics Consortium**

Naomi R Wray* 1, 2

Stephan Ripke* 3, 4, 5

Manuel Mattheisen* 6, 7, 8

Maciej Trzaskowski 1

Enda M Byrne 1

Abdel Abdellaoui 9

Mark J Adams 10

Esben Agerbo 11, 12, 13

Tracy M Air 14

Till F M Andlauer 15, 16

Silviu-Alin Bacanu 17

Marie Bækvad-Hansen 13, 18

Aartjan T F Beekman 19

Tim B Bigdeli 17, 20

Elisabeth B Binder 15, 21

Julien Bryois 22

Henriette N Buttenschøn 13, 23, 24

Jonas Bybjerg-Grauholm 13, 18

Na Cai 25, 26

Enrique Castelao 27

Jane Hvarregaard Christensen 8, 13, 24

Toni-Kim Clarke 10

Jonathan R I Coleman 28

Lucía Colodro-Conde 29

Baptiste Couvy-Duchesne 2, 30

Nick Craddock 31

Gregory E Crawford 32, 33

Gail Davies 34

Franziska Degenhardt 35

Eske M Derks 29

Nese Direk 36, 37

Conor V Dolan 9

Erin C Dunn 38, 39, 40

Thalia C Eley 28

Valentina Escott-Price 41

Farnush Farhadi Hassan Kiadeh 42

Hilary K Finucane 43, 44

Jerome C Foo 45

Andreas J Forstner 35, 46, 47, 48

Josef Frank 45

Héléna A Gaspar 28

Michael Gill 49

Fernando S Goes 50

Scott D Gordon 29

Jakob Grove 8, 13, 24, 51

Lynsey S Hall 10, 52

Christine Søholm Hansen 13, 18

Thomas F Hansen 53, 54, 55

Stefan Herms 35, 47

Ian B Hickie 56

Per Hoffmann 35, 47

Georg Homuth 57

Carsten Horn 58

Jouke-Jan Hottenga 9

David M Hougaard 13, 18

David M Howard 10, 28

Marcus Ising 59

Rick Jansen 19

Ian Jones 60

Lisa A Jones 61

Eric Jorgenson 62

James A Knowles 63

Isaac S Kohane 64, 65, 66

Julia Kraft 4

Warren W. Kretzschmar 67

Zoltán Kutalik 68, 69

Yihan Li 67

Penelope A Lind 29

Donald J MacIntyre 70, 71

Dean F MacKinnon 50

Robert M Maier 2

Wolfgang Maier 72

Jonathan Marchini 73

Hamdi Mbarek 9

Patrick McGrath 74

Peter McGuffin 28

Sarah E Medland 29

Divya Mehta 2, 75

Christel M Middeldorp 9, 76, 77

Evelin Mihailov 78

Yuri Milaneschi 19

Lili Milani 78

Francis M Mondimore 50

Grant W Montgomery 1

Sara Mostafavi 79, 80

Niamh Mullins 28

Matthias Nauck 81, 82

Bernard Ng 80

Michel G Nivard 9

Dale R Nyholt 83

Paul F O'Reilly 28

Hogni Oskarsson 84

Michael J Owen 60

Jodie N Painter 29

Carsten Bøcker Pedersen 11, 12, 13

Marianne Giørtz Pedersen 11, 12, 13

Roseann E Peterson 17, 85

Wouter J Peyrot 19

Giorgio Pistis 27

Danielle Posthuma 86, 87

Jorge A Quiroz 88

Per Qvist 8, 13, 24

John P Rice 89

Brien P. Riley 17

Margarita Rivera 28, 90

Saira Saeed Mirza 36

Robert Schoevers 91

Eva C Schulte 92, 93

Ling Shen 62

Jianxin Shi 94

Stanley I Shyn 95

Engilbert Sigurdsson 96

Grant C B Sinnamon 97

Johannes H Smit 19

Daniel J Smith 98

Hreinn Stefansson 99

Stacy Steinberg 99

Fabian Streit 45

Jana Strohmaier 45

Katherine E Tansey 100

Henning Teismann 101

Alexander Teumer 102

Wesley Thompson 13, 54, 103, 104

Pippa A Thomson 105

Thorgeir E Thorgeirsson 99

Matthew Traylor 106

Jens Treutlein 45

Vassily Trubetskoy 4

André G Uitterlinden 107

Daniel Umbricht 108

Sandra Van der Auwera 109

Albert M van Hemert 110

Alexander Viktorin 22

Peter M Visscher 1, 2

Yunpeng Wang 13, 54, 104

Bradley T. Webb 111

Shantel Marie Weinsheimer 13, 54

Jürgen Wellmann 101

Gonneke Willemsen 9

Stephanie H Witt 45

Yang Wu 1

Hualin S Xi 112

Jian Yang 2, 113

Futao Zhang 1

Volker Arolt 114

Bernhard T Baune 114, 115, 116

Klaus Berger 101

Dorret I Boomsma 9

Sven Cichon 35, 47, 117, 118

Udo Dannlowski 114

EJC de Geus 9, 119

J Raymond DePaulo 50

Enrico Domenici 120

Katharina Domschke 121, 122

Tõnu Esko 5, 78

Hans J Grabe 109

Steven P Hamilton 123

Caroline Hayward 124

Andrew C Heath 89

Kenneth S Kendler 17

Stefan Kloiber 59, 125, 126

Glyn Lewis 127

Qingqin S Li 128

Susanne Lucae 59

Pamela AF Madden 89

Patrik K Magnusson 22

Nicholas G Martin 29

Andrew M McIntosh 10, 34

Andres Metspalu 78, 129

Ole Mors 13, 130

Preben Bo Mortensen 11, 12, 13, 24

Bertram Müller-Myhsok 15, 131, 132

Merete Nordentoft 13, 133

Markus M Nöthen 35

Michael C O'Donovan 60

Sara A Paciga 134

Nancy L Pedersen 22

Brenda WJH Penninx 19

Roy H Perlis 38, 135

David J Porteous 105

James B Potash 136

Martin Preisig 27

Marcella Rietschel 45

Catherine Schaefer 62

Thomas G Schulze 45, 93, 137, 138, 139

Jordan W Smoller 38, 39, 40

Kari Stefansson 99, 140

Henning Tiemeier 36, 141, 142

Rudolf Uher 143

Henry Völzke 102

Myrna M Weissman 74, 144

Thomas Werge 13, 54, 145

Jaako Kaprio 146

Cathryn M Lewis* 28, 147

Douglas F Levinson* 148

Gerome Breen* 28, 149

Anders D Børglum* 8, 13, 24

Patrick F Sullivan* 22, 150, 151

1, Institute for Molecular Bioscience, The University of Queensland, Brisbane, QLD, AU

2, Queensland Brain Institute, The University of Queensland, Brisbane, QLD, AU

3, Analytic and Translational Genetics Unit, Massachusetts General Hospital, Boston, MA, US

4, Department of Psychiatry and Psychotherapy, Universitätsmedizin Berlin Campus Charité Mitte, Berlin, DE

5, Medical and Population Genetics, Broad Institute, Cambridge, MA, US

6, Department of Psychiatry, Psychosomatics and Psychotherapy, University of Wurzburg, Wurzburg, DE

7, Centre for Psychiatry Research, Department of Clinical Neuroscience, Karolinska Institutet, Stockholm, SE

8, Department of Biomedicine, Aarhus University, Aarhus, DK

9, Dept of Biological Psychology & EMGO+ Institute for Health and Care Research, Vrije Universiteit Amsterdam, Amsterdam, NL

10, Division of Psychiatry, University of Edinburgh, Edinburgh, GB

11, Centre for Integrated Register-based Research, Aarhus University, Aarhus, DK

12, National Centre for Register-Based Research, Aarhus University, Aarhus, DK

13, iPSYCH, The Lundbeck Foundation Initiative for Integrative Psychiatric Research,, DK

14, Discipline of Psychiatry, University of Adelaide, Adelaide, SA, AU

15, Department of Translational Research in Psychiatry, Max Planck Institute of Psychiatry, Munich, DE

16, Department of Neurology, Klinikum rechts der Isar, Technical University of Munich, Munich, DE

17, Department of Psychiatry, Virginia Commonwealth University, Richmond, VA, US

18, Center for Neonatal Screening, Department for Congenital Disorders, Statens Serum Institut, Copenhagen, DK

19, Department of Psychiatry, Vrije Universiteit Medical Center and GGZ inGeest, Amsterdam, NL

20, Virginia Institute for Psychiatric and Behavior Genetics, Richmond, VA, US

21, Department of Psychiatry and Behavioral Sciences, Emory University School of Medicine, Atlanta, GA, US

22, Department of Medical Epidemiology and Biostatistics, Karolinska Institutet, Stockholm, SE

23, Department of Clinical Medicine, Translational Neuropsychiatry Unit, Aarhus University, Aarhus, DK

24, iSEQ, Centre for Integrative Sequencing, Aarhus University, Aarhus, DK

25, Human Genetics, Wellcome Trust Sanger Institute, Cambridge, GB

26, Statistical genomics and systems genetics, European Bioinformatics Institute (EMBL-EBI), Cambridge, GB

27, Department of Psychiatry, Lausanne University Hospital and University of Lausanne, Lausanne, CH

28, Social, Genetic and Developmental Psychiatry Centre, King's College London, London, GB

29, Genetics and Computational Biology, QIMR Berghofer Medical Research Institute, Brisbane, QLD, AU

30, Centre for Advanced Imaging, The University of Queensland, Brisbane, QLD, AU

31, Psychological Medicine, Cardiff University, Cardiff, GB

32, Center for Genomic and Computational Biology, Duke University, Durham, NC, US

33, Department of Pediatrics, Division of Medical Genetics, Duke University, Durham, NC, US

34, Centre for Cognitive Ageing and Cognitive Epidemiology, University of Edinburgh, Edinburgh, GB

35, Institute of Human Genetics, University of Bonn, School of Medicine & University Hospital Bonn, Bonn, DE

36, Epidemiology, Erasmus MC, Rotterdam, Zuid-Holland, NL

37, Psychiatry, Dokuz Eylul University School Of Medicine, Izmir, TR

38, Department of Psychiatry, Massachusetts General Hospital, Boston, MA, US

39, Psychiatric and Neurodevelopmental Genetics Unit (PNGU), Massachusetts General Hospital, Boston, MA, US

40, Stanley Center for Psychiatric Research, Broad Institute, Cambridge, MA, US

41, Neuroscience and Mental Health, Cardiff University, Cardiff, GB

42, Bioinformatics, University of British Columbia, Vancouver, BC, CA

43, Department of Epidemiology, Harvard T.H. Chan School of Public Health, Boston, MA, US

44, Department of Mathematics, Massachusetts Institute of Technology, Cambridge, MA, US

45, Department of Genetic Epidemiology in Psychiatry, Central Institute of Mental Health,  Medical Faculty Mannheim, Heidelberg University, Mannheim, Baden-Württemberg, DE

46, Department of Psychiatry (UPK), University of Basel, Basel, CH

47, Department of Biomedicine, University of Basel, Basel, CH

48, Centre for Human Genetics, University of Marburg, Marburg, DE

49, Department of Psychiatry, Trinity College Dublin, Dublin, IE

50, Psychiatry & Behavioral Sciences, Johns Hopkins University, Baltimore, MD, US

51, Bioinformatics Research Centre, Aarhus University, Aarhus, DK

52, Institute of Genetic Medicine, Newcastle University, Newcastle upon Tyne, GB

53, Danish Headache Centre, Department of Neurology, Rigshospitalet, Glostrup, DK

54, Institute of Biological Psychiatry, Mental Health Center Sct. Hans, Mental Health Services Capital Region of Denmark, Copenhagen, DK

55, iPSYCH, The Lundbeck Foundation Initiative for Psychiatric Research, Copenhagen, DK

56, Brain and Mind Centre, University of Sydney, Sydney, NSW, AU

57, Interfaculty Institute for Genetics and Functional Genomics, Department of Functional Genomics, University Medicine and Ernst Moritz Arndt University Greifswald, Greifswald, Mecklenburg-Vorpommern, DE

58, Roche Pharmaceutical Research and Early Development, Pharmaceutical Sciences, Roche Innovation Center Basel, F. Hoffmann-La Roche Ltd, Basel, CH

59, Max Planck Institute of Psychiatry, Munich, DE

60, MRC Centre for Neuropsychiatric Genetics and Genomics, Cardiff University, Cardiff, GB

61, Department of Psychological Medicine, University of Worcester, Worcester, GB

62, Division of Research, Kaiser Permanente Northern California, Oakland, CA, US

63, Psychiatry & The Behavioral Sciences, University of Southern California, Los Angeles, CA, US

64, Department of Biomedical Informatics, Harvard Medical School, Boston, MA, US

65, Department of Medicine, Brigham and Women's Hospital, Boston, MA, US

66, Informatics Program, Boston Children's Hospital, Boston, MA, US

67, Wellcome Trust Centre for Human Genetics, University of Oxford, Oxford, GB

68, Institute of Social and Preventive Medicine (IUMSP), Lausanne University Hospital and University of Lausanne, Lausanne, VD, CH

69, Swiss Institute of Bioinformatics, Lausanne, VD, CH

70, Division of Psychiatry, Centre for Clinical Brain Sciences, University of Edinburgh, Edinburgh, GB

71, Mental Health, NHS 24, Glasgow, GB

72, Department of Psychiatry and Psychotherapy, University of Bonn, Bonn, DE

73, Statistics, University of Oxford, Oxford, GB

74, Psychiatry, Columbia University College of Physicians and Surgeons, New York, NY, US

75, School of Psychology and Counseling, Queensland University of Technology, Brisbane, QLD, AU

76, Child and Youth Mental Health Service, Children's Health Queensland Hospital and Health Service, South Brisbane, QLD, AU

77, Child Health Research Centre, University of Queensland, Brisbane, QLD, AU

78, Estonian Genome Center, University of Tartu, Tartu, EE

79, Medical Genetics, University of British Columbia, Vancouver, BC, CA

80, Statistics, University of British Columbia, Vancouver, BC, CA

81, DZHK (German Centre for Cardiovascular Research), Partner Site Greifswald, University Medicine, University Medicine Greifswald, Greifswald, Mecklenburg-Vorpommern, DE

82, Institute of Clinical Chemistry and Laboratory Medicine, University Medicine Greifswald, Greifswald, Mecklenburg-Vorpommern, DE

83, Institute of Health and Biomedical Innovation, Queensland University of Technology, Brisbane, QLD, AU

84, Humus, Reykjavik, IS

85, Virginia Institute for Psychiatric & Behavioral Genetics, Virginia Commonwealth University, Richmond, VA, US

86, Clinical Genetics, Vrije Universiteit Medical Center, Amsterdam, NL

87, Complex Trait Genetics, Vrije Universiteit Amsterdam, Amsterdam, NL

88, Solid Biosciences, Boston, MA, US

89, Department of Psychiatry, Washington University in Saint Louis School of Medicine, Saint Louis, MO, US

90, Department of Biochemistry and Molecular Biology II, Institute of Neurosciences, Biomedical Research Center (CIBM), University of Granada, Granada, ES

91, Department of Psychiatry, University of Groningen, University Medical Center Groningen, Groningen, NL

92, Department of Psychiatry and Psychotherapy, University Hospital, Ludwig Maximilian University Munich, Munich, DE

93, Institute of Psychiatric Phenomics and Genomics (IPPG), University Hospital, Ludwig Maximilian University Munich, Munich, DE

94, Division of Cancer Epidemiology and Genetics, National Cancer Institute, Bethesda, MD, US

95, Behavioral Health Services, Kaiser Permanente Washington, Seattle, WA, US

96, Faculty of Medicine, Department of Psychiatry, University of Iceland, Reykjavik, IS

97, School of Medicine and Dentistry, James Cook University, Townsville, QLD, AU

98, Institute of Health and Wellbeing, University of Glasgow, Glasgow, GB

99, deCODE Genetics / Amgen, Reykjavik, IS

100, College of Biomedical and Life Sciences, Cardiff University, Cardiff, GB

101, Institute of Epidemiology and Social Medicine, University of Münster, Münster, Nordrhein-Westfalen, DE

102, Institute for Community Medicine, University Medicine Greifswald, Greifswald, Mecklenburg-Vorpommern, DE

103, Department of Psychiatry, University of California, San Diego, San Diego, CA, US

104, KG Jebsen Centre for Psychosis Research, Norway Division of Mental Health and Addiction, Oslo University Hospital, Oslo, NO

105, Medical Genetics Section, CGEM, IGMM, University of Edinburgh, Edinburgh, GB

106, Clinical Neurosciences, University of Cambridge, Cambridge, GB

107, Internal Medicine, Erasmus MC, Rotterdam, Zuid-Holland, NL

108, Roche Pharmaceutical Research and Early Development, Neuroscience, Ophthalmology and Rare Diseases Discovery & Translational Medicine Area, Roche Innovation Center Basel, F. Hoffmann-La Roche Ltd, Basel, CH

109, Department of Psychiatry and Psychotherapy, University Medicine Greifswald, Greifswald, Mecklenburg-Vorpommern, DE

110, Department of Psychiatry, Leiden University Medical Center, Leiden, NL

111, Virginia Institute for Psychiatric & Behavioral Genetics, Virginia Commonwealth University, Richmond, VA, US

112, Computational Sciences Center of Emphasis, Pfizer Global Research and Development, Cambridge, MA, US

113, Institute for Molecular Bioscience; Queensland Brain Institute, The University of Queensland, Brisbane, QLD, AU

114, Department of Psychiatry, University of Münster, Münster, Nordrhein-Westfalen, DE

115, Department of Psychiatry, Melbourne Medical School, University of Melbourne, Melbourne, AU

116, Florey Institute for Neuroscience and Mental Health, University of Melbourne, Melbourne, AU

117, Institute of Medical Genetics and Pathology, University Hospital Basel, University of Basel, Basel, CH

118, Institute of Neuroscience and Medicine (INM-1), Research Center Juelich, Juelich, DE

119, Amsterdam Public Health Institute, Vrije Universiteit Medical Center, Amsterdam, NL

120, Centre for Integrative Biology, Università degli Studi di Trento, Trento, Trentino-Alto Adige, IT

121, Department of Psychiatry and Psychotherapy, Medical Center - University of Freiburg, Faculty of Medicine, University of Freiburg, Freiburg, DE

122, Center for NeuroModulation, Faculty of Medicine, University of Freiburg, Freiburg, DE

123, Psychiatry, Kaiser Permanente Northern California, San Francisco, CA, US

124, Medical Research Council Human Genetics Unit, Institute of Genetics and Molecular Medicine, University of Edinburgh, Edinburgh, GB

125, Department of Psychiatry, University of Toronto, Toronto, ON, CA

126, Centre for Addiction and Mental Health, Toronto, ON, CA

127, Division of Psychiatry, University College London, London, GB

128, Neuroscience Therapeutic Area, Janssen Research and Development, LLC, Titusville, NJ, US

129, Institute of Molecular and Cell Biology, University of Tartu, Tartu, EE

130, Psychosis Research Unit, Aarhus University Hospital, Risskov, Aarhus, DK

131, Munich Cluster for Systems Neurology (SyNergy), Munich, DE

132, University of Liverpool, Liverpool, GB

133, Mental Health Center Copenhagen, Copenhagen Universtity Hospital, Copenhagen, DK

134, Human Genetics and Computational Biomedicine, Pfizer Global Research and Development, Groton, CT, US

135, Psychiatry, Harvard Medical School, Boston, MA, US

136, Psychiatry, University of Iowa, Iowa City, IA, US

137, Department of Psychiatry and Behavioral Sciences, Johns Hopkins University, Baltimore, MD, US

138, Department of Psychiatry and Psychotherapy, University Medical Center Göttingen, Goettingen, Niedersachsen, DE

139, Human Genetics Branch, NIMH Division of Intramural Research Programs, Bethesda, MD, US

140, Faculty of Medicine, University of Iceland, Reykjavik, IS

141, Child and Adolescent Psychiatry, Erasmus MC, Rotterdam, Zuid-Holland, NL

142, Psychiatry, Erasmus MC, Rotterdam, Zuid-Holland, NL

143, Psychiatry, Dalhousie University, Halifax, NS, CA

144, Division of Translational Epidemiology, New York State Psychiatric Institute, New York, NY, US

145, Department of Clinical Medicine, University of Copenhagen, Copenhagen, DK

146, Institute for Molecular Medicine Finland FIMM, Helsinki Institute of Life Science, University of Helsinki, FI

147, Department of Medical & Molecular Genetics, King's College London, London, GB

148, Psychiatry & Behavioral Sciences, Stanford University, Stanford, CA, US

149, NIHR Maudsley Biomedical Research Centre, King's College London, London, GB

150, Genetics, University of North Carolina at Chapel Hill, Chapel Hill, NC, US

151, Psychiatry, University of North Carolina at Chapel Hill, Chapel Hill, NC, US
